# Comparing cannabinoid extracts for treating cancer-related symptoms: a randomized placebo-controlled, triple-blind aggregate n-of-1 clinical trial

**DOI:** 10.64898/2026.05.31.26354558

**Authors:** Philippa H. Hawley, Kevin Wade, Paul Daeninck, Ed Fitzgibbon, Marc Kerba, Craig E. Goldie, Ilana Kopolovic

**Affiliations:** BC Cancer Vancouver Centre; University of British Columbia, Department of Medicine, Division of Palliative Care; BC Cancer, University of British Columbia, Division of Palliative Care; Dept of Internal Medicine, Max Rady College of Medicine, University of Manitoba, Winnipeg Regional Palliative Care Program; The Ottawa Hospital Cancer Center. University of Ottawa, Ottawa Hospital Research Institute; Department of Oncology, Division of Radiation Oncology, Arthur Child Comprehensive Cancer Centre, University of Calgary; Queen’s University, Department of Medicine, Division of Palliative Care, Kingston Health Sciences Center, Providence Care Hospital; Durham Regional Cancer Centre, Lakeridge Health, Department of Medicine, Division of Oncology, Queen’s University, University of Toronto

**Author notes:** **Corresponding Author:** Mailing Address, Dr P. Hawley, Pain & Symptom Management/Palliative Care (4^th^ floor), BC Cancer-Vancouver Centre, 600 West 10^th^ Ave., Vancouver, British Columbia, V5Z 4E6, Canada, [No X handle]. **Co-author list:** Please note that the Investigator from Montreal, Dr. Michael Dworkind, has retired and is not able to be a co-author. He is acknowledged.

**Keywords:** Cancer, symptoms, medical cannabis, cannabinoids, crossover study

## Abstract

**Context:** Despite widespread use of medical cannabinoids for cancer-related symptom management, systematic reviews consistently call for more clinical trial evidence.

**Objectives:** This study aimed to determine and explore responses to medical cannabis extracts for cancer-related symptoms using patient-centred methodology.

**Methods:** An aggregate N-of-1 study of clinically stable but symptomatic outpatients from 8 Canadian cancer centres, comparing three blinded sublingual extracts (THC; CBD; 1:1) with placebo, self-titrated within a prescribed schedule for four consecutive days each in randomized sequence for up to three cycles (total 16-48 days). The primary outcome was the frequency of at least a 1.4-point (20%) improvement in a 7-point Patient Global Impression of Change (PGIC) for at least one extract over placebo.

**Results:** The primary outcome was achieved in 50/89 (56%) participants (p<0.001), with no significant preference of one extract over another on average, but a clear preference between extracts for most individuals. Changes in a modified Edmonton Symptom Assessment score and participant preference (n=91) confirmed these findings. Improved sleep, tiredness and anxiety contributed most to the overall improvement regardless of primary symptom. There were no demographic predictors of response. Mild adverse effects were common with all extracts including placebo but resolved rapidly on dose reduction/cessation. Moderate/severe adverse effects were rare but associated with THC.

**Conclusions:** Medical cannabis extracts can be meaningfully beneficial for cancer-related symptoms in approximately 50% of patients, particularly for sleep and related symptoms. A starting dose of 2.5mg of THC/CBD three times a day was well-tolerated. Personalization of treatment is required to optimize response.

**Key Messages:** Three cannabinoid extracts (THC; CBD; and 1:1) were significantly more effective than placebo based on a Patient Global Impression of Change, a modified Edmonton Symptom Assessment System and participant preference. The most helpful extract differed between individuals. Benefits were mostly in sleep, anxiety, and daytime tiredness irrespective of primary symptom.

## Introduction

Medical cannabis extracts are consumed by many people living with or after cancer for symptom management at all stages of the disease although there is limited randomized clinical trial evidence informing the use of these extracts (1-4). There has been limited acknowledgement of the potential for benefit from medical cannabis products amongst oncologists and other physicians, and stigmatization is still widespread. This can lead to conflict between patients and clinicians. There have been many calls for clinical trials to provide evidence-based clinical guidance (5-7).

There is substantial diversity between individuals in endocannabinoid physiology (8). Widespread anecdotal evidence suggests that some patients find cannabinoids very helpful, and others do not. Trial designs that don’t accommodate patient heterogeneity may underestimate the benefits experienced by some participants (9,10.) Parallel group studies have difficulty recruiting with a placebo-only arm, and advanced cancer patients have a high dropout rate due to clinical instability. Symptom severity scores alone are limited in detecting clinically meaningful changes (11).

Aggregate n-of-1 trials are multi-cycle, placebo-controlled, short crossover trials, with all participants receiving all interventions, serving as their own controls. Clinical stability is essential for differences between treatment effects to be attributed to the intervention and not altered disease status. This design provides the strongest possible evidence about the efficacy of a treatment in an individual (12). A series (aggregate) of such trials can yield an estimate of the population effect comparable to parallel group randomized controlled trials with far fewer participants, by removing the need for baseline matching of treatment arms and avoidance of potential confounders (13,14). Without the need for matching of treatment arms, recruitment can be demographically diverse, maximizing generalizability of conclusions.

## Methods

### Study Design

This multi-center Canadian study consisted of a series of 91 individual triple-blind n-of-1 trials comparing three different types of cannabinoid oil extracts with placebo oils. The study was designed according to the Consolidated Standards of Reporting Trials (CONSORT) statement and CONSORT Extension for n-of-1 trials.

### Participants

Eligible patients were ≥19 years old, competent, clinically stable, with a life expectancy ≥4 months, ECOG ≤2, with at least one of three target symptoms rated ≥4/10 related to living with (or justifiably fearing) cancer of any kind, at any stage, or its treatment. Their most troublesome symptom had to be pain, sleep disturbance or anxiety. Recruitment was to three corresponding subgroups equally. Symptoms unrelated to cancer were excluded. A very high hereditary cancer risk was eligible in the anxiety or sleep disturbance arms. Patients were excluded if they had a cannabis use disorder (CUDIT-R score ≥8) (15), other substance use disorder, recent cannabis consumption, pregnancy, breastfeeding or reproductive potential, a concurrent relevant clinical trial, serious cardiovascular disease, or having a personal or first degree relative with schizophrenia. Participants agreed to alcohol and driving restrictions during participation. Patients stable on daily oral or intermittent parenteral systemic therapies with no immediate side-effects or need for anticipatory medication were allowed, including immunotherapies (16).

### Recruitment

Recruitment was through seven symptom management/palliative care clinics in Canadian provincial cancer centres and a medical cannabis clinic closely associated with one (Montreal). Eligible patients self-identified from REB-approved advertising or were identified by medical staff and contacted (with permission) by research staff to obtain informed consent.

### Study Products

Medical cannabis extracts were tetrahydrocannabinol (THC) and cannabidiol (CBD) and a 1:1 mix of the two. Placebo was the same vehicle oil (olive or mixed-chain triglycerides) as the active extracts. Three different suppliers of similar research-grade extracts were required over the course of the study, all certified by Health Canada for research purposes according to Good Manufacturing Practices (GMP), and all products were low or very low in terpene content. Rosemary or peppermint was added if necessary for optimal blinding.

### Randomization and Masking

Extract sequence was randomized according to a pre-coded computer-generated randomization schedule for each patient, coordinated by the Vancouver Study Site Pharmacist. Participants were told that flavoring may be added to mask the taste of any oils, so would not assume that flavoured extracts were placebo. Colored syringes and opaque containers ensured visual blinding.

### Study Procedures

After consenting and baseline assessments, four 30-ml bottles labelled from 1 to 4 were given/sent to participants. The number did not correspond to the same extract for each participant. Each treatment period was 4 days, minimum participation 1 cycle (16 days), up to 3 cycles allowed. Participants were unblinded after return of documentation and remaining extracts. Given the short onset and limited duration of effects following sublingual administration, the first 2 days of each 4-day period were considered the washout period to allow for resolution of effects from the prior extract. A treatment-free washout would have made participation too long for maintenance of clinical stability and compromised compliance with the study routine. A treatment pause between extracts was allowed for potentially confounding events, e.g., vaccination, or for unexpected international travel, providing clinical stability was not compromised.

Participants were asked to take a minimum of 0.15ml (3 drops) per day, and additionally up to 0.15 ml every 4 hours as needed on an escalating schedule, for a total of no more than 15 drops (0.75ml) per day in the first cycle. In cycles 2 and 3 they could start at the dose worked up to by the end of the first cycle for that extract and could increase further to a maximum of 18 drops (0.9ml) per day. Cannabinoid concentrations were THC: 20.1-27.0mg/ml (0% CBD), CBD: 19.4-27.5mg/ml (0.5-1.4mg/ml THC), and 1:1: 8.0-18.1mg/ml THC and 10.7-16.0mg/ml CBD. The maximum amount of THC and CBD that this represents from each of the three suppliers varied from 18.0 to 24.6mg/day.

Compliance and adverse event documentation was facilitated by study personnel telephoning at least every 4 days, actively soliciting reports of effects and adverse events. An investigator call/visit took place at the end of each cycle. No non-study medications were allowed to be changed throughout the study. Use of pre-established breakthrough/rescue medications were documented.

The study was performed in line with the principles of the Declaration of Helsinki, approved by Health Canada and applicable local Research Ethics Boards. An independent Data and Safety Monitoring Committee reviewed the study at pre-specified recruitment targets. The trial was registered at clinicaltrials.gov, NCT03948074.

### Outcome Assessment

The primary outcome was the difference in average Patient Global Impression of Change (PGIC) score across the two non-washout days of each treatment cycle from baseline for each oil as compared to placebo. Placebo (and nocebo) effects were expected. A PGIC provides a single, general estimate of improvement or deterioration. The PGIC scale we used asked subjects to rate their status as compared with baseline as:

1. No change (or condition worse),
2. Almost the same (hardly any change at all),
3. A little better (no noticeable change),
4. Somewhat better (change has not made any real difference,
5. Moderately better (slight but noticeable change),
6. Better (definite improvement that has made a real and worthwhile difference),
7. A great deal better and a considerable improvement that has made all the difference.

We considered a 1.4-point (20%) improvement in PGIC score over the individual placebo response to be the Minimal Clinically Important Difference (MCID). This was derived from data from five non-cancer pain cohorts which found that change greater than 1.35 points on an 11-point scale was clinically important. Head-to-head comparison of a 7-point scale with a 15-point scale had found no significant difference in performance (17).

The secondary outcome was improvement above placebo in a modified version of the revised Edmonton Symptom Assessment System (ESAS-r) including sleep disturbance and night sweats (18). The ESAS-r-SN was completed daily in the morning for the sleep arm, at bedtime otherwise.

An additional outcome was patient oil preference stated before unblinding.

Exploratory outcomes included optimized doses in participants’ last cycle; breakdown in symptom severity ratings as a proportion of overall ESAS-r-SN change; adverse event reports; and analysis of demographic variables with respect to response rates.

### Statistical considerations

Sample size was estimated by conducting seven power simulations comparing each treatment period via the open-source software R version 3.6.1 on a 64-bit Windows platform (19). Each scenario assumed that at least one of the three active treatments offered an improvement in PGIC of 1.4 points over response to placebo. This generated a target of 30 participants per subgroup, plus 25% added to allow for expected attrition, i.e., 113 total. Pooled analyses of the PGIC and ESAS-r-SN were conducted using a linear mixed effects model, allowing for fixed effects of treatment cycle, treatment, index symptom and the interaction between treatment and index symptom, and for a random subject effect.

## Results

Total recruitment time was 43 months, between February 2021 and July 2025, with two breaks for approval of new study product. See Fig. 1 CONSORT diagram.

**Figure 1.**
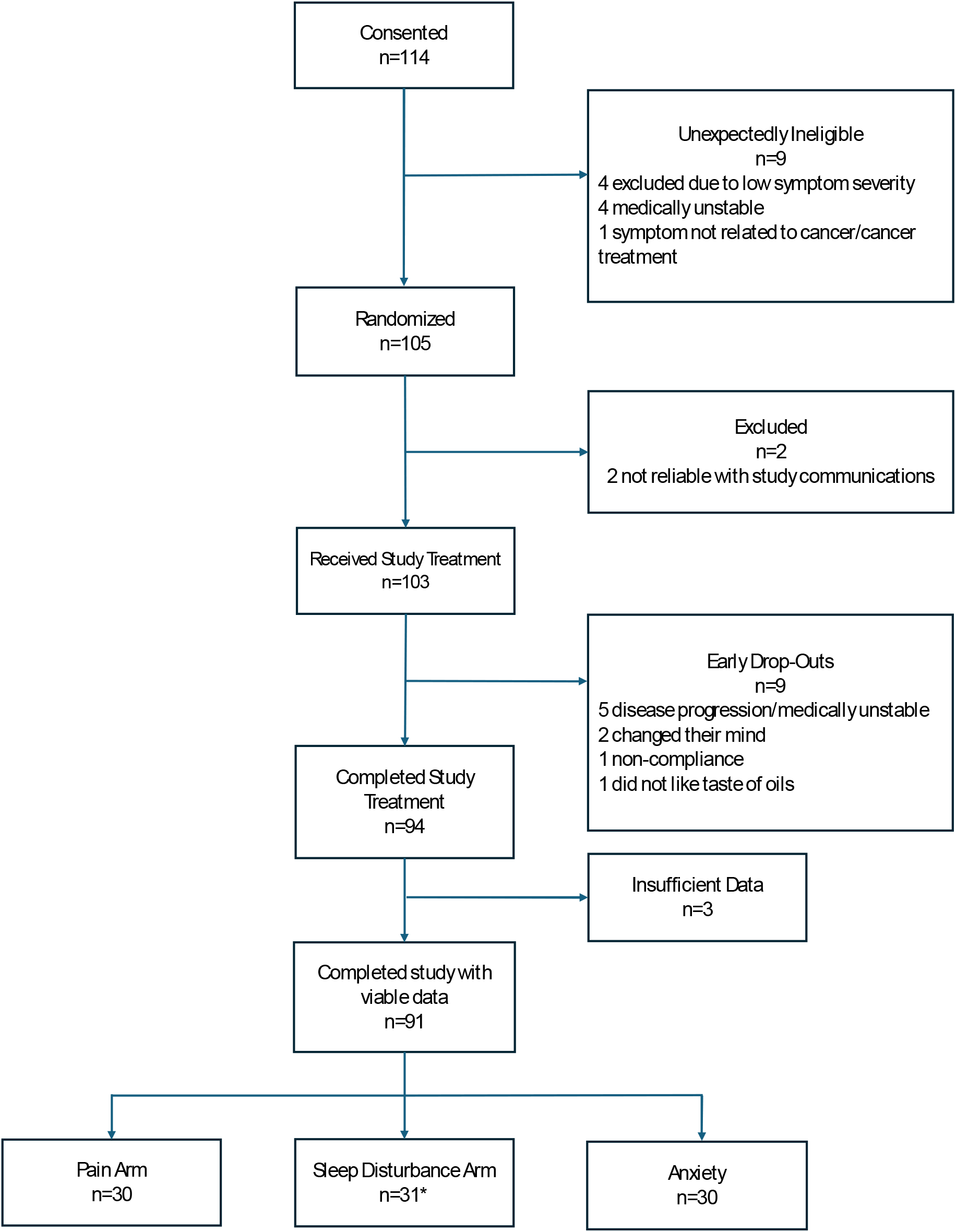
CONSORT Diagram

### Demographics

The age of participants ranged from 27 to 86 yrs., mean 60, median 62. The anxiety subgroup was younger (mean 55). One participant (anxiety) had a very high hereditary cancer risk but had not yet had cancer. Breast cancer was the most common primary cancer in all three subgroups. All stages of cancer were represented: 20% no active disease, 30% stages 1-3, and 36% stage 4. “Unknowns” (14%) were mostly haematologic cancers where the staging system does not apply. Baseline ESAS-r-SN scores ranged from 4 to 73/110, mean 36. The mean baseline ESAS-r (excluding sleep and night sweats) was 28/90. For details see supplemental baseline demographics file with embedded figures and tables.

### Response Rates

Figure 2(a) shows the forest plot for differences in PGIC from placebo (represented by the continuous vertical line), for each individual participant averaged from the last 2 days of each treatment period. Each participant has three symbols, indicating response to THC (squares), CBD (circles), and 1:1 (triangles). The dotted vertical line indicates a 1.4-point (20%) improvement in PGIC above that participant’s placebo response, which was on average 2.17 points. Where there was individual regression analysis significance in that participant’s n-of-1 study the symbols are black, otherwise grey. Individual logistic regression was only possible in those that completed 3 cycles, so lack of individual trial statistical significance does not imply lack of clinically meaningful change. A star indicates completion of one cycle only.

**Figure 2.**
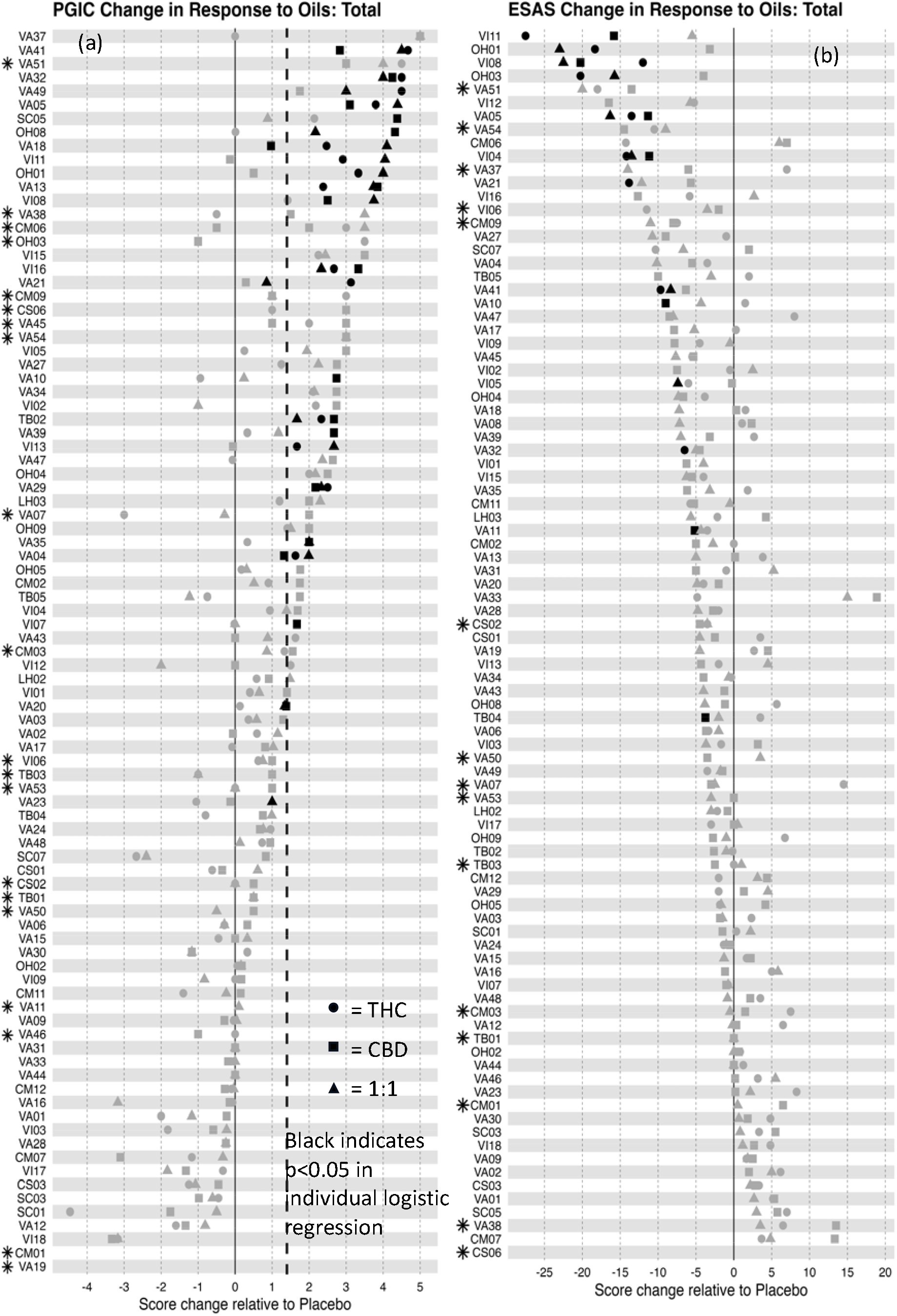
PGIC (a) and ESAS-r-SN (b) change relative to placebo for all n-of-1 trials, * indicates only one cycle completed. See supplemental Figure A for ESAS-r-SN in same order as PGIC.

Fifty (56%) of the 89 participants with complete PGIC data reported at least a1.4-point improvement compared to placebo with at least one extract. Subgroup analysis showed response rates of 50% for the pain, 47% for sleep, and 60% for anxiety subgroup. Though there was often a marked difference in response within individuals, each active extract was associated with the best and worst responses in approximately equal numbers of participants.

The pooled PGIC analysis (Table 1) showed that the mean additional changes in PGIC above the placebo response was strongly statistically significant for all three extracts (p<0.001 or p=0.001), but not significantly different in effectiveness from each other on average. Compared with those who only completed one cycle (17/89), participants completing two or three cycles did not show a significant additional increase in PGIC scores, indicating that one cycle was sufficient for inclusion in the pooled analysis.

**Table 1.**
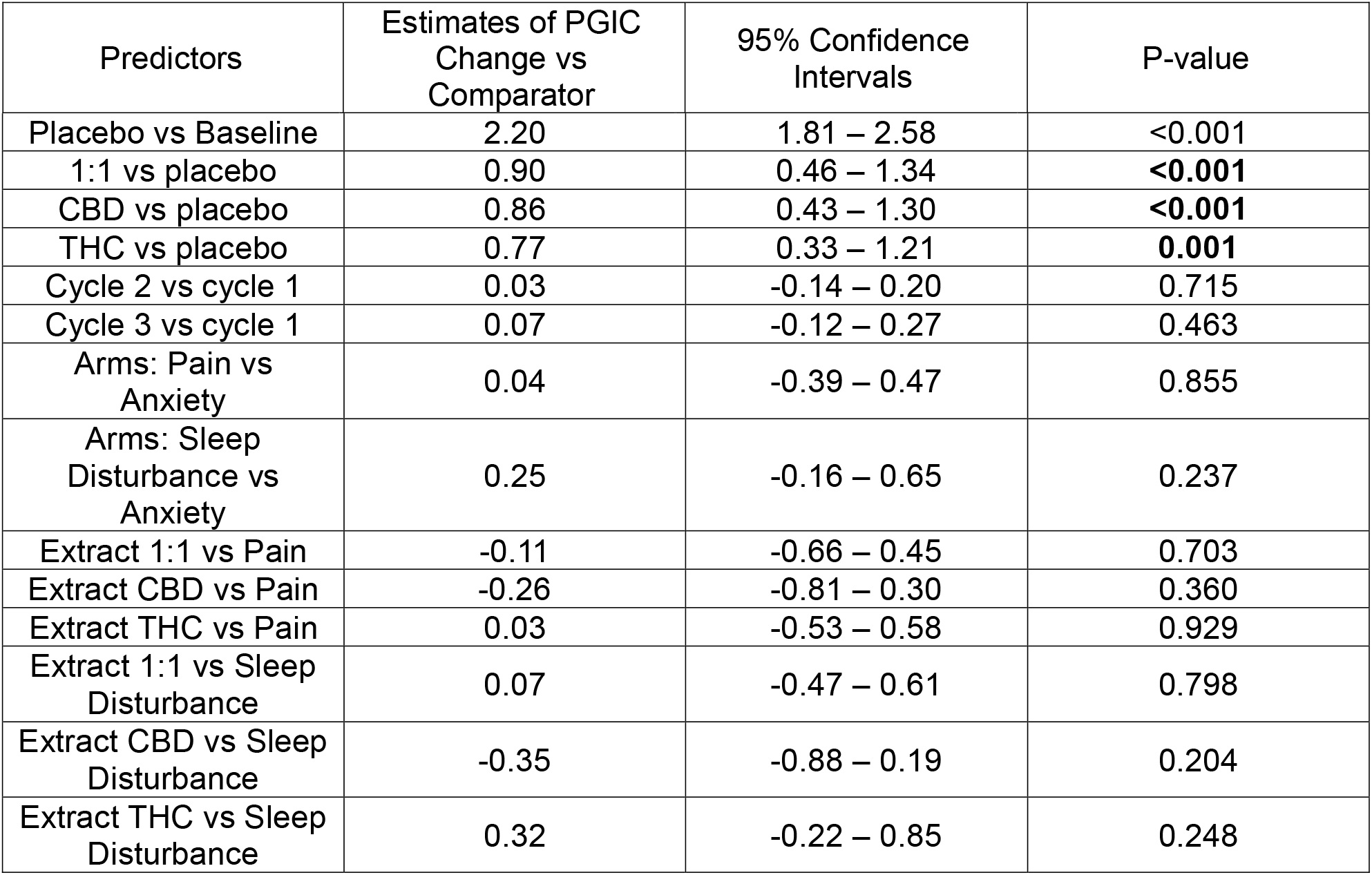
Pooled PGIC Analysis (n=89)

| Predictors | Estimates of PGIC<br>Change vs<br>Comparator | 95% Confidence<br>Intervals | P-value |
| --- | --- | --- | --- |
| Placebo vs Baseline | 2.20 | 1.81 – 2.58 | <0.001 |
| 1:1 vs placebo | 0.90 | 0.46 – 1.34 | **<0.001** |
| CBD vs placebo | 0.86 | 0.43 – 1.30 | **<0.001** |
| THC vs placebo | 0.77 | 0.33 – 1.21 | **0.001** |
| Cycle 2 vs cycle 1 | 0.03 | -0.14 – 0.20 | 0.715 |
| Cycle 3 vs cycle 1 | 0.07 | -0.12 – 0.27 | 0.463 |
| Arms: Pain vs Anxiety | 0.04 | -0.39 – 0.47 | 0.855 |
| Arms: Sleep Disturbance vs Anxiety | 0.25 | -0.16 – 0.65 | 0.237 |
| Extract 1:1 vs Pain | -0.11 | -0.66 – 0.45 | 0.703 |
| Extract CBD vs Pain | -0.26 | -0.81 – 0.30 | 0.360 |
| Extract THC vs Pain | 0.03 | -0.53 – 0.58 | 0.929 |
| Extract 1:1 vs Sleep Disturbance | 0.07 | -0.47 – 0.61 | 0.798 |
| Extract CBD vs Sleep Disturbance | -0.35 | -0.88 – 0.19 | 0.204 |
| Extract THC vs Sleep Disturbance | 0.32 | -0.22 – 0.85 | 0.248 |

Figure 2(b) shows the ESAS-r-SN results from all 91 participants in order of descending magnitude of best benefit (see supplemental Figure 5 for ESAS-r-SN results shown in the same order as in the PGIC plot).

The pooled total ESAS-r-SN analysis (Table 2) shows a statistically significant overall benefit from all three extracts (p<0.001-0.005), with the sleep subgroup contributing most. The response of the anxiety subgroup increased more relative to the first cycle (p<0.001) than for the PGIC. Participants in this subgroup were understandably slower to increase their doses than the pain/sleep subgroups.

**Table 2.**
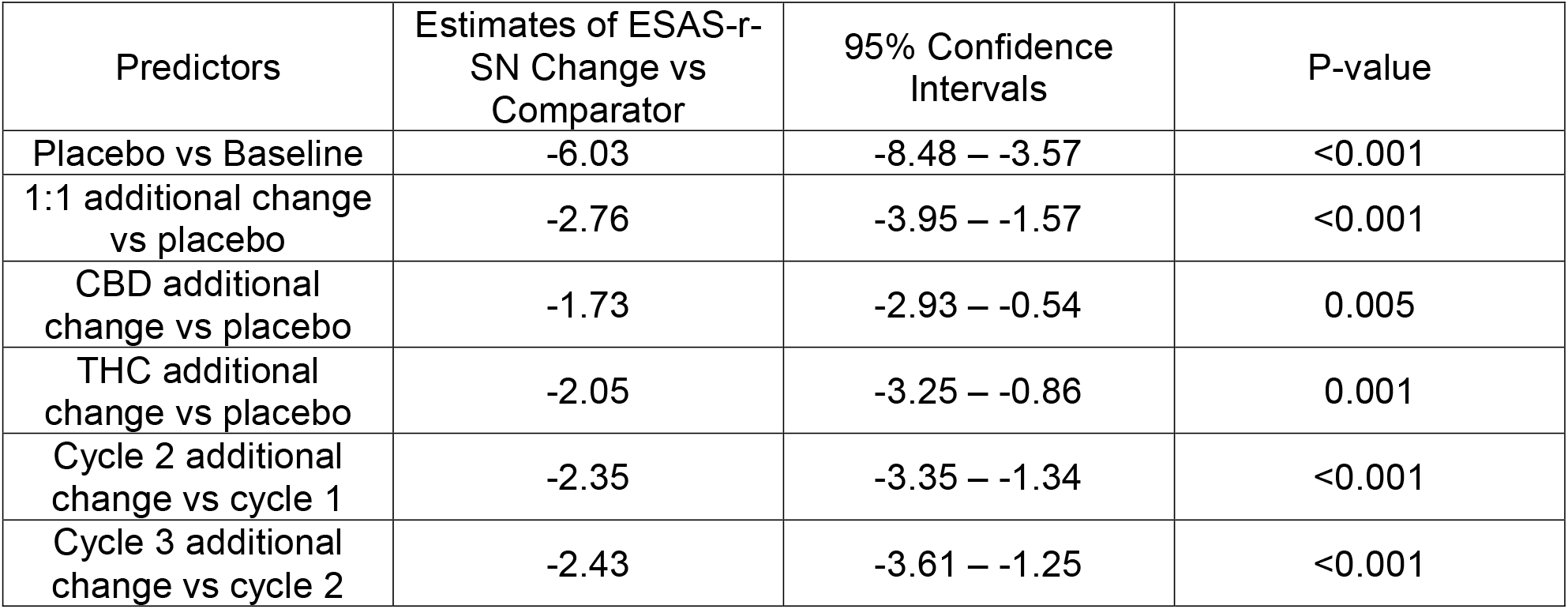
Pooled ESAS-r-SN analysis.

The even spread of blinded participant’s stated active oil preference further confirmed the PGIC and ESAS findings, see Fig. 3.

**Figure 3.**
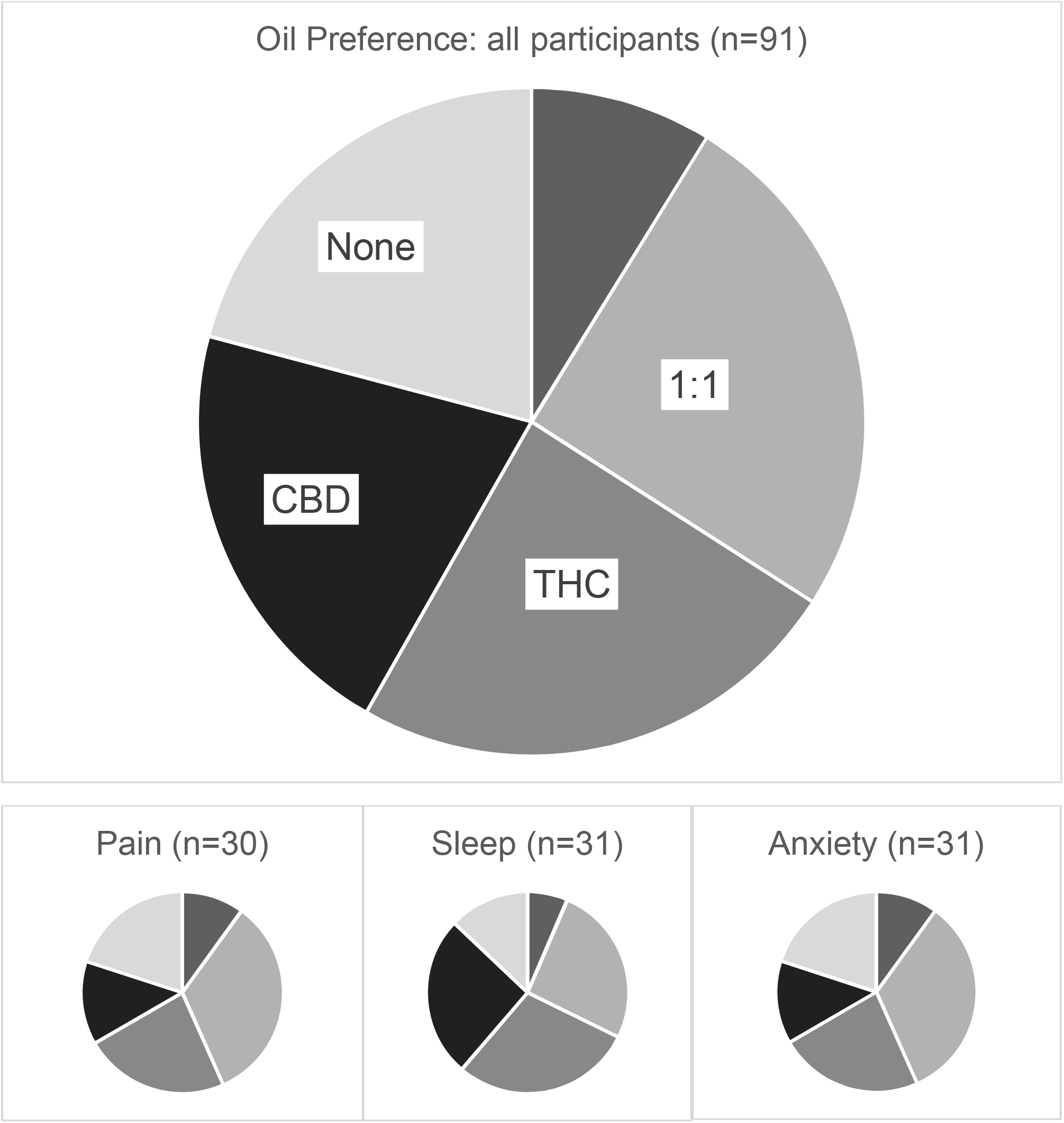
Blinded extract preference Shading and order of each extract in all charts is the same. Unlabelled “slice” is placebo.

### Doses

There was a trend for lower optimized doses in the anxiety subgroup, but dosing was otherwise independent of symptom number or intensity at baseline.

The mean total cannabinoid doses taken on the last day of the last treatment period for each extract were very similar: THC 9.64mg; CBD 10.47mg and 1:1 8.56mg (Table 3). The final average total cannabinoid dose of 1:1 was close to that of THC and CBD alone, which suggests that CBD has independent and additive effects to THC, not just reducing THC’s side-effects.

**Table 3.**
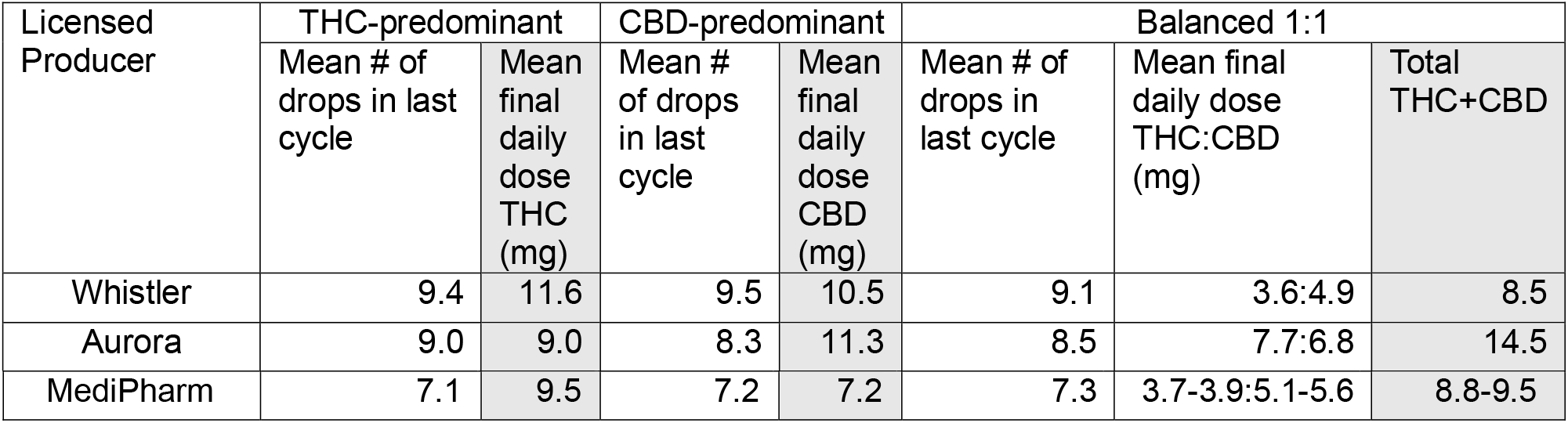
Mean doses of cannabinoids taken on final day of each extract.

### Relative Contribution of Each Symptom to Overall Response

The mean and median total baseline ESAS-r-SN scores were 36/110, ranging from 4 to 73, with mean individual symptom severity worst for sleep disturbance (5.8), tiredness (5.4), pain (4.2), wellbeing (4.2), drowsiness (4.1), anxiety (3.8), depression (2.3), night sweats (2.2), shortness of breath (1.6), loss of appetite (1.5) and nausea (0.8). For comparison with other study purposes, the mean total ESAS-r was 28/90. There was no significant difference in mean total ESAS-r-SN between subgroups.

As shown in Fig. 4, improvements in sleep, anxiety and tiredness contributed most to the total ESAS-r-SN changes, but there were differences between subgroups. Breathlessness only featured along with anxiety. The improvements seen in anxiety and tiredness in the pain subgroup were greater compared to other symptoms. In the sleep subgroup, sleep was the biggest contributor to benefit but still only contributed a quarter of the overall benefit. The anxiety and sleep subgroups showed improvement in night sweats. Improvement in appetite, overall wellbeing and depression were not major contributors, and nausea was only mildly relevant in the pain subgroup, reflecting low baseline prevalence.

### Adverse Events

As expected, most participants reported multiple mild or moderate adverse effects with multiple extracts and placebo (Table 4). Most were seen with higher doses; however, all resolved rapidly with dose reduction or cessation. There were 31 adverse events that were moderate or severe and deemed “possibly” or “probably” related to the intervention by the investigator. Amongst these, only THC was found to increase the odds of an adverse event, by 189% vs placebo (Supplemental Table 6). Two participants experienced dizziness/fall, one worst when on the 1:1 oil and one worst on CBD. Another (in the anxiety subgroup) withdrew due to multiple anxiety-related symptoms experienced whilst on all extracts, including placebo. These participants completed at least one cycle and all adverse events are included in the analysis. Reasons for dropping out prior to completion of the minimum one cycle required for analysis (9/103) were not due to adverse events but were for unexpected change in circumstances, inability to comply with the study protocol, and one disliking the taste of at least one extract.

**Table 4.**
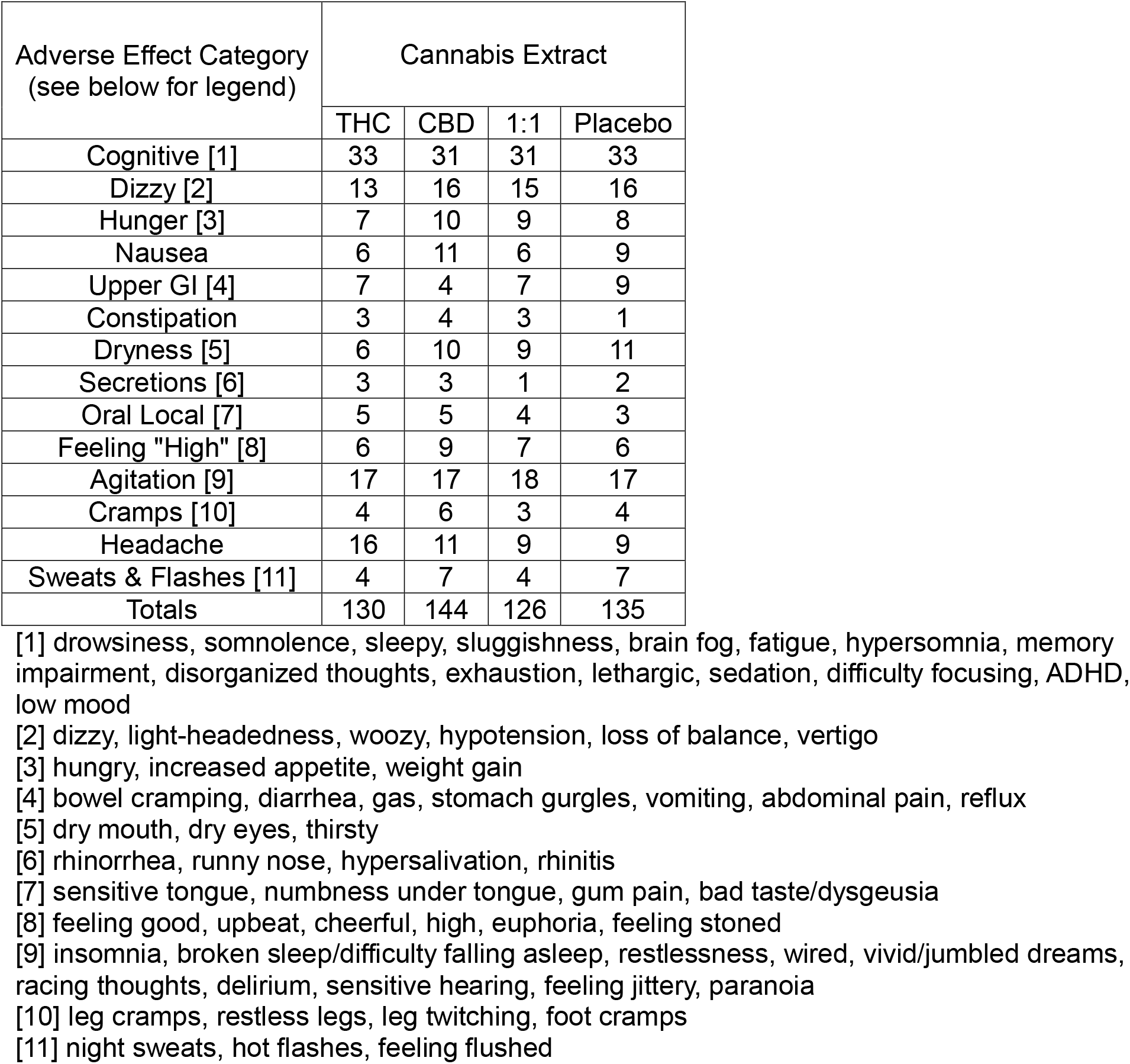
All severities of adverse events according to extract treatment period (n=91) Occurrence of adverse effect occurring at any point in individuals’ participation

### Demographic Predictors of Response

Though the responders were on average slightly younger than the non-responders, this difference was not statistically significant and there was no association between response and sex, primary cancer type or stage (supplemental Table 6).

## Discussion

We found that more than half of participants experienced a clinically meaningful benefit over placebo with at least one type of cannabis extract, highly statistically significant in the pooled analyses. Although no extract type was routinely better than the others on average, most participants had a clearly better response to one type than the others, most likely reflecting heterogeneity in participant endocannabinoid tone. Participants were able to self-titrate appropriately and there were few moderate or severe side effects reported, all of which resolved rapidly on dose reduction/cessation. These results are very similar to a 34-participant non-cancer chronic pain n-of-1 series reported in 2004 (20).

In contrast to most prior studies, by using an aggregate n-of-1 design, we were able to isolate the effects of the different active oils from baseline treatment arm differences with an efficient sample size and include a diverse spread of the population. By keeping participation short and requiring clinical stability for eligibility we were able to avoid confounding interventions and complications. These design features facilitated identification of highly statistical differences in multiple outcomes measured in three different ways. Ensuring that each subject personally benefited from their participation also created a sense of personal investment and added meaning to their illness.

Our threshold for definition of clinically meaningful response may have under-represented the real response rate. Clear preference for an active oil was higher (66%) than the PGIC-derived response rate (56%), suggesting that our threshold for definition of response of 1.4 points better than placebo may have under-reported the minimal clinically important benefit. Additionally, some participants reporting no preference were unable to choose between more than one active oil, but more than one was perceived as better than placebo, indicating likelihood of further under-identification of potentially relevant benefit.

Our finding that no preparation was better than the others on average, but most participants found one to provide greater benefit individually may further explain why trials that focus on a single preparation may have underestimated the potential benefit of cannabinoids. Restricting patients to a single extract ignores the known heterogeneity in endocannabinoid physiology. A recent meta-analysis of clinical trials of cannabinoids describes the perils of over-simplification of complex physiology and pharmacology and conclude that patient-reported outcomes, particularly global impressions of change, indicate that a real therapeutic effect is likely (21). Our >50% clinical response rate is consistent with patient surveys and the few open-label studies where patients were allowed to personalize their cannabinoid therapy (22-24). Canadian cancer survivors have been found to choose a THC-predominant product 36% of the time, CBD-predominant 23% of the time, and a 1:1 mix 33% of the time, a high proportion reporting sleep and anxiety as the primary motivation for consumption (2). As with systemic oncology treatments, though it would be convenient to be able to recommend a single agent for everyone, this would clearly not meet the needs of a diverse population. Until we have cannabinoid response predictive testing analogous to mutation detection in tumours, personalized cannabinoid therapy is required. Clinical trials should allow for personalization of therapy, for example, having n-of-1 trials to identify preferred extract type as a run-in for long-term studies.

The threshold for MCID in the total ESAS-r-SN has implication for future research. As a post-hoc analysis, taking the PGIC response as the “gold standard” indicator, a 56% response rate would indicate that a point change of -4 points would be the MCID (figure 2(c)).^18^ However, when the two forest plots are placed side by side with participants in the same order (Figure 2(b) it can be appreciated that they are not entirely consistent. Patients may assign differing levels of importance to changes in different symptoms. By transferring each subgroup’s PGIC-determined response rate to the ESAS-r-SN forest plots, we found the MCID to be -3 in the pain subgroup, -4 in the anxiety subgroup, and -7 in the sleep disturbance subgroup.

Though THC-only products were associated with moderate/severe side-effects more often than the CBD or balanced oils and placebo, a quarter of participants still preferred a THC-predominant oil over the others, inferring that the benefits for many participants outweighed the side-effects.

This study provides an evidence-based starting protocol for therapeutic trials of medical cannabis extracts in stable adult cancer patients, at all stages of disease, irrespective of sex. A starting dose of 2.5mg total THC/CBD self-titrated up to every 4 hrs to best balance between benefits and side-effects would be well-tolerated by most patients, similar to the recommendations of a 2021 non-cancer pain management consensus recommendation (25).

### Strengths and Limitations

Though the total duration of treatment was short (4-12 days total per extract), this ensured that clinical stability was maintained, and potentially confounding treatments were avoided, thereby maximizing the validity of comparisons between treatment periods. There was no treatment-free period between extracts but the 2-day washout between data inclusion periods was more than adequate to avoid carryover effects.

The PGIC scale did not include gradations of worsening, which may have led to a skewing of the results to positivity, however “no change or worsening” was an option, and any worsening of symptoms was captured by the ESAS-r-SN, which confirmed the findings, also supported by majority patient preference for extracts over placebo.

Half the participants were recruited through one centre (Vancouver), however there was no difference in response rate at that site as compared with the other sites, and all centres provided comparable services.

Very few participants were previously experienced with cannabinoids. This facilitated effective blinding, but doses may need to be higher in patients who are tolerant to the effects of cannabinoids.

That three different study product suppliers were required to complete the study could be considered a limitation of the study in terms of reproducibility, however, there was no difference in outcomes between product suppliers, suggesting that the findings can be generalized to similar products available in Canada and other regulated jurisdictions, e.g., US, Australia, UK, ensuring immediate clinical relevance. The transmucosal route of administration was chosen to avoid potential harms from inhalation and allow for accurate dosing, and rapid titration, and carries a lower risk of accidental ingestion by children and pets than edibles (26,27).

## Supporting information

Baseline demographics

PGIC and ESASrSN in same order

Related adverse events

Demographic logistic regression

## Data Availability

All data produced in the present study are available upon reasonable request to the authors

## Disclosures and Acknowledgments

### Authorship Statement

Philippa Hawley was the sponsor and principal investigator (Vancouver site), responsible for study conceptualization and design, protocol development, Health Canada communication (including writing the Investigator Brochure), study implementation, financing, compliance with regulators, holding the Cannabis Research License, analysing and interpreting the data, writing and editing the manuscript and preparing the attachments for publication. Paul Daeninck and Ilana Kopolovic validated the original data, were site investigators, and contributed to review and editing of the manuscript The other authors were site investigators and contributed to the review and editing of the manuscript.

### AI disclosure

Artificial intelligence was not used at any point in the preparation of this manuscript.

## List of Tables and Figures

Editor, please note that all figures are formatted for black and white/greyscale to ensure readability when photocopied. Colour files for figures 2-4 are available if needed.

**Figure 4.**
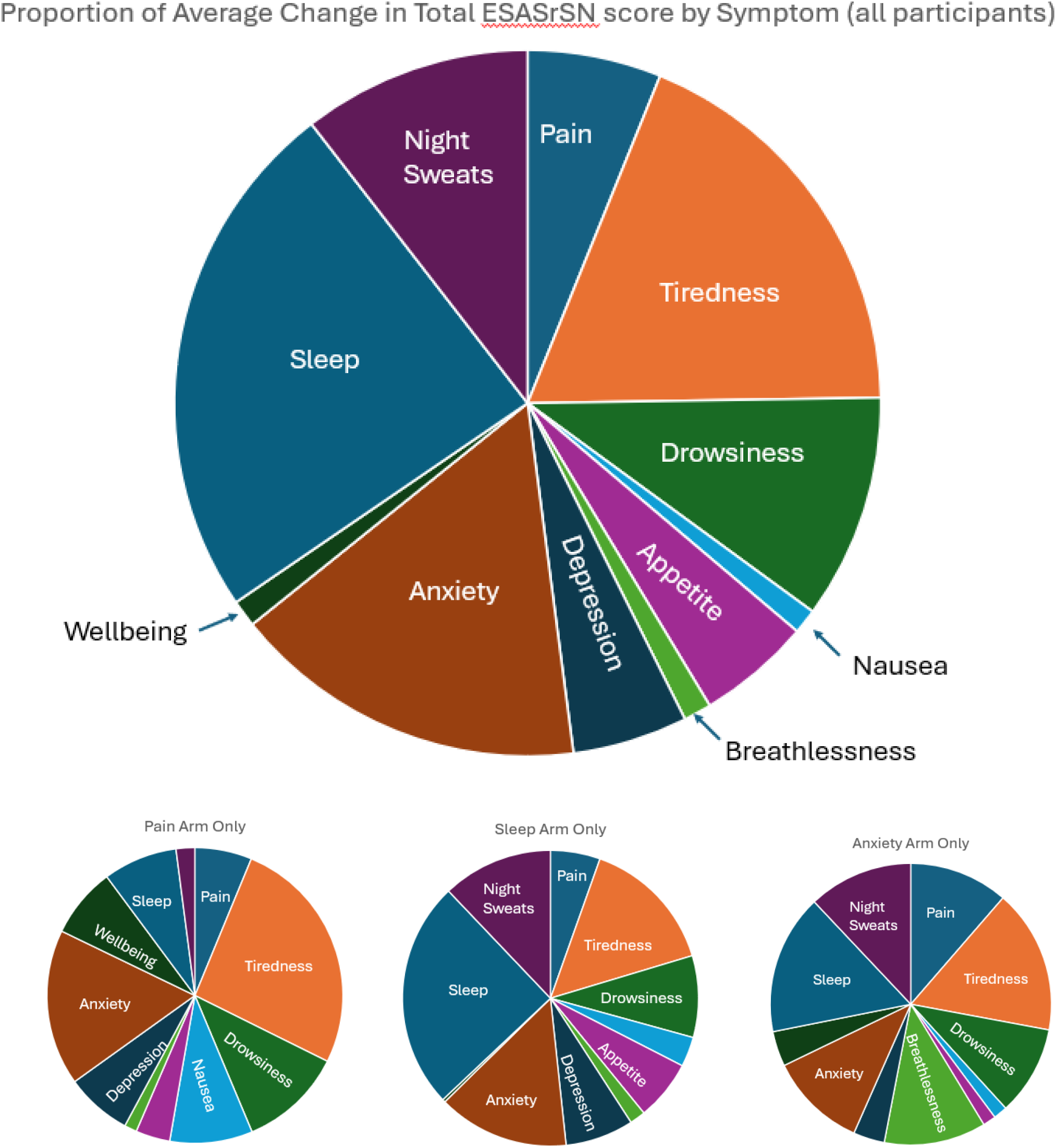
Proportion of Average Change in Total ESASrSN score by Symptom (all participants)

### Main manuscript

Table 1 Pooled PGIC analysis

Table 2 Pooled ESAS-R-SN analysis

Table 3 Mean doses of cannabinoids taken on final day of each extract

Table 4 Adverse events according to extract treatment period

Fig 1 Consort diagram

Fig 2 Forest plot of all individual n-of-1 studies

Fig 3 Blinded extract preference

Fig 4 Contributions of each symptom to overall benefit

### Supplemental Files

Baseline demographics (Excel file with multiple embedded charts)

Table 5. Logistic regression for risk of moderate/severe adverse events deemed possibly or probably related to each extract

Table 6. Demographic logistic regression

## Supplemental Tables

**Supplemental Table S5.**
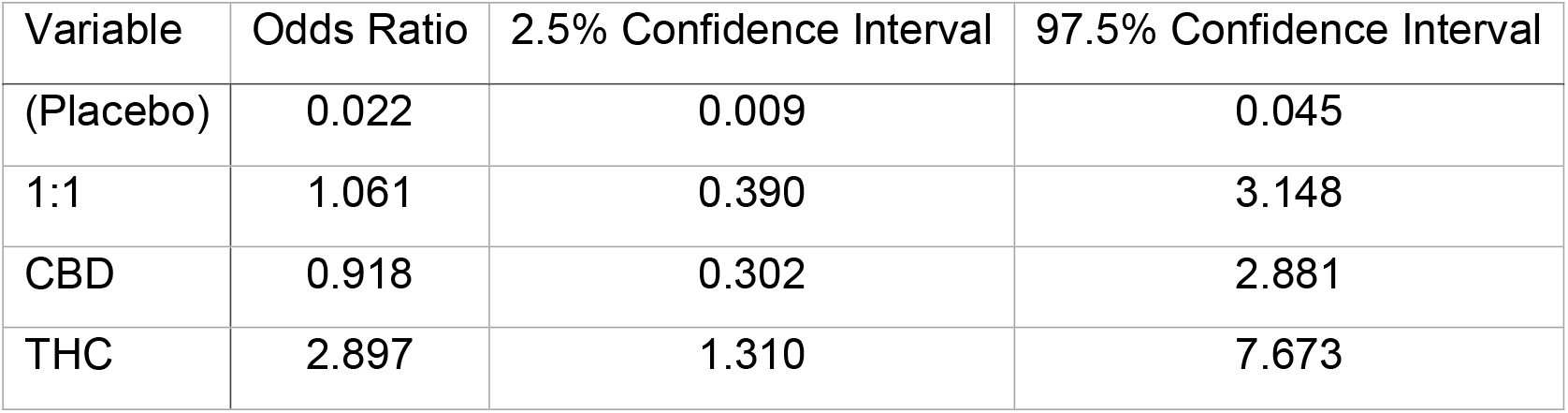
Logistic regression for risk of moderate/severe adverse events deemed possibly or probably related to each extract.

**Supplemental Table S6.**
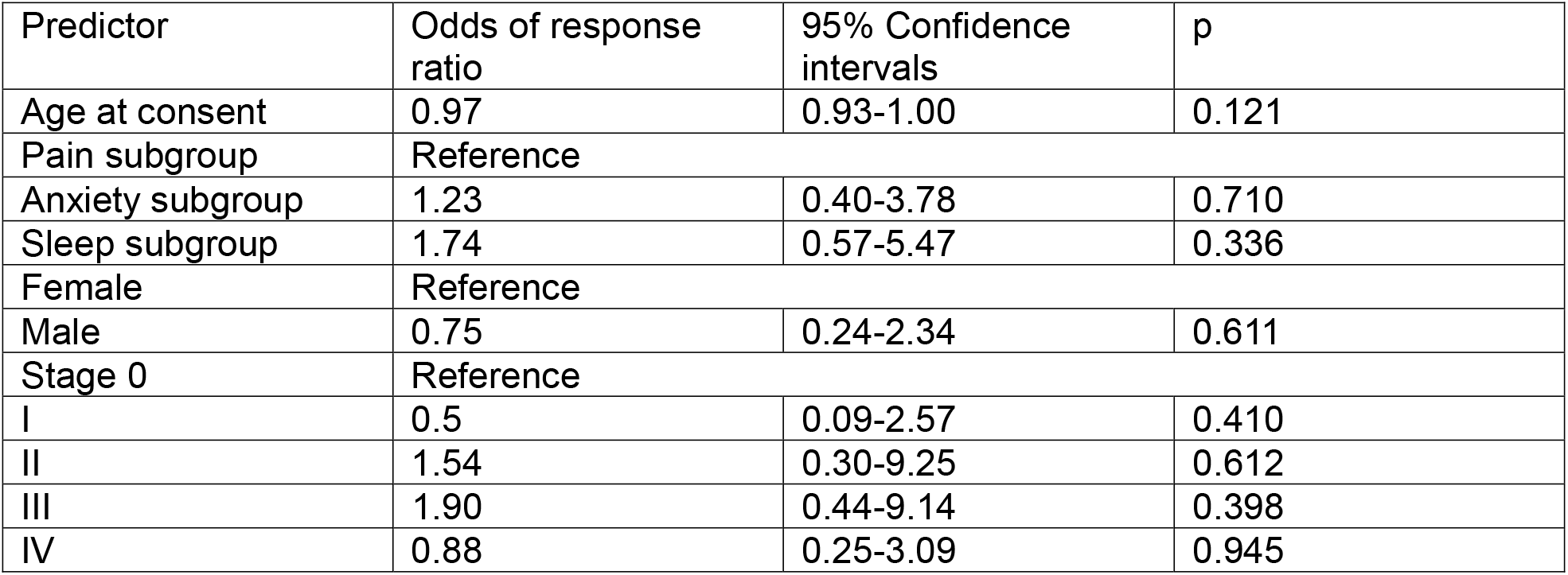
Demographics logistic regression (Obs=84, R^2^Tjur 0.0870)

## Acknowledgements

1. Dr. Michael Dworkind (now retired), for being the Sante Cannabis site lead (Investigator) in Montreal.
2. Isabella Ghement for statistical analysis plan development, and Mark Pitblado for providing the interim and final statistical reports.
3. The Pan-Canadian Palliative Care Research Collaboration, particularly Dr. James Downar (Ottawa), for assistance with coordination of protocol review, recruiting participating sites, and for assistance with editing.
4. MediPharm (Ontario), for making suitable study product available for purchase at short notice.

## References

1. Newcomer KF, Barnell EK, Zhang F et al: Surgical cancer patients’ attitudes and beliefs about cannabis at a midwest comprehensive cancer center. Surgical Oncology Insight, 2025,2(3):100154. https://www.surgoncinsight.org/article/S2950-2470(25)00033-7/fulltext

2. Lee RM, Donnan J, Harris N et al: A cross-sectional survey of the prevalence and patterns of using cannabis as a sleep aid in Canadian cancer survivors. J Cancer Surviv 2025 Feb;19(1):386–396. https://pubmed.ncbi.nlm.nih.gov/37837502/

3. Hawley P, Gobbo M, Afghari N. The impact of legalization of access to recreational Cannabis on Canadian medical users with Cancer. BMC Health Serv Res. 2020 Oct 27;20(1):977. https://pmc.ncbi.nlm.nih.gov/articles/PMC7590602/

4. Braun IM, Bohlke K, Abrams DI et al: Cannabis and Cannabinoids in Adults with Cancer: ASCO Guideline. J Clin Oncol. 2024 May 1;42(13):1575–1593. https://pubmed.ncbi.nlm.nih.gov/38478773/

5. Cyr C, Arboleda MF, Aggarwal SK et al: Cannabis in palliative care: current challenges and practical recommendations. Ann Palliat Med. 2018 Oct;7(4):463-477. Erratum in: Ann Palliat Med. 2019 Apr;8(2):215–217. https://pubmed.ncbi.nlm.nih.gov/30180728/

6. Braun IM, Wright A, Peteet J, et al: Medical Oncologists’ Beliefs, Practices, and Knowledge Regarding Marijuana Used Therapeutically: A Nationally Representative Survey Study. J Clin Oncol 2018;36:1957–1962. https://pubmed.ncbi.nlm.nih.gov/29746226/

7. Allan GM, Ramji J, Perry D, et al: Simplified guideline for prescribing medical cannabinoids in primary care. Can Fam Physician. 2018 Feb;64(2):111–120. https://pubmed.ncbi.nlm.nih.gov/29449241/

8. Boehnke KF, J. Scott R, Litinas E et al: Cannabis Use Preferences and Decision-making Among a Cross-sectional Cohort of Medical Cannabis Patients with Chronic Pain, The Journal of Pain 2019, 20(11):1362–1372. https://pubmed.ncbi.nlm.nih.gov/31132510/

9. Hardy J, Greer R, Huggett G et al: Phase IIb Randomized, Placebo-Controlled, Dose-Escalating, Double-Blind Study of Cannabidiol Oil for the Relief of Symptoms in Advanced Cancer (MedCan1-CBD). J Clin Oncol. 2023 Mar 1;41(7):1444–1452. https://pubmed.ncbi.nlm.nih.gov/36409969/

10. Hardy JR, Greer RM, Pelecanos AM et al: Medicinal cannabis for symptom control in advanced cancer: a double-blind, placebo-controlled, randomised clinical trial of 1:1 tetrahydrocannabinol and cannabidiol. Support Care Cancer. 2025 Jul 24;33(8):715. https://pubmed.ncbi.nlm.nih.gov/40705150/

11. Hui D, Bruera E. The Edmonton Symptom Assessment System 25 Years Later: Past, Present, and Future Developments. J Pain Symptom Manage. 2017 Mar;53(3):630–643. https://pubmed.ncbi.nlm.nih.gov/28042071/

12. Mitchell GK. Generating high-quality clinical evidence in palliative care using N-of-1 Trials. J Pall Med 2010, 13(10):1185–6. https://pubmed.ncbi.nlm.nih.gov/20942759/

13. Lillie EO, Patay B, Diamant J et al: The n-of-1 clinical trial: the ultimate strategy for individualizing medicine? Per Med. 2011 Mar;8(2):161–173. https://pubmed.ncbi.nlm.nih.gov/21695041/

14. Hui D, Zhukovsky DS, Bruera E. Which treatment is better? Ascertaining patient preferences with crossover randomized controlled trials. J Pain Symptom Manage. 2015 Mar;49(3):625–31. https://pubmed.ncbi.nlm.nih.gov/25555446/

15. Adamson SJ, Kay-Lambkin FJ, Baker AL et al: An improved brief measure of cannabis misuse: the Cannabis Use Disorders Identification Test-Revised (CUDIT-R). Drug Alcohol Depend. 2010 Jul 1;110(1-2):137–43. https://pubmed.ncbi.nlm.nih.gov/20347232/

16. Waissengrin B, Leshem Y, Taya M, et al: The use of medical cannabis concomitantly with immune checkpoint inhibitors in non-small cell lung cancer: A sigh of relief? European Journal of Cancer 2023, 180:52–61. https://pubmed.ncbi.nlm.nih.gov/36535195/

17. Kamper SJ, Maher CG, Mackay G. Global rating of change scales: a review of strengths and weaknesses and considerations for design. J Man Manip Ther. 2009;17(3):163–70. https://pubmed.ncbi.nlm.nih.gov/20046623/

18. Hui D, Shamieh O, Paiva CE et al: Minimal clinically important differences in the Edmonton Symptom Assessment Scale in cancer patients: A prospective, multicenter study. Cancer. 2015 Sep 1;121(17):3027–35. https://pubmed.ncbi.nlm.nih.gov/26059846/

19. Senn S. Sample size considerations for n-of-1 trials. Stat Methods Med Res. 2019 Feb;28(2):372–383. https://pubmed.ncbi.nlm.nih.gov/28882093/

20. Notcutt W, Price M, Miller R et al. Initial experiences with medicinal extracts of cannabis for chronic pain results from 34 ‘n of 1’ studies. Anaesthesia 2004, 59(5):440–452. https://pubmed.ncbi.nlm.nih.gov/15096238/

21. Castle R, Marzolf J, Morris M, Bushell W. Meta-analysis of medical cannabis outcomes and associations with cancer. Front Oncol 2025; 15:1–23. 10.3389/fonc.2025.1490621

22. Ghasemiesfe M, Ravi D, Vali M et al: Marijuana use, respiratory symptoms, and pulmonary function: a systematic review and meta-analysis. Ann Intern Med. 2018; 169(2):106–115. https://pubmed.ncbi.nlm.nih.gov/29971337/

23. Myran DT, Tanuseputro P, Auger N et al: Edible Cannabis Legalization and Unintentional Poisonings in Children. N Engl J Med. 2022 Aug 25;387(8):757–759. https://pubmed.ncbi.nlm.nih.gov/36001718/

24. Waissengrin B, Urban D, Leshem Y et al: Patterns of use of medical cannabis among Israeli cancer patients: a single institution experience. J Pain Symptom Manage. 2015 Feb;49(2):223–30. https://pubmed.ncbi.nlm.nih.gov/24937161/

25. Bhaskar A, Bell A, Boivin M et al: Consensus recommendations on dosing and administration of medical cannabis to treat chronic pain: results of a modified Delphi process. J Cannabis Res. 2021 Jul 2;3(1):22. https://pmc.ncbi.nlm.nih.gov/articles/PMC8252988/

26. Tramèr MR, Carroll D, Campbell FA et al: Cannabinoids for control of chemotherapy induced nausea and vomiting: quantitative systematic review. BMJ. 2001 Jul 7;323(7303):16–21. https://pubmed.ncbi.nlm.nih.gov/11440936/

27. Bar-Lev Schleider L, Raphael Mechoulam R, Violeta Lederman V, et al: Prospective analysis of safety and efficacy of medical cannabis in large, unselected population of patients with cancer, European Journal of Internal Medicine 2018, 49:37–43. https://pubmed.ncbi.nlm.nih.gov/29482741/

