## Supplementary material for "Comparing cannabinoid extracts for treating cancer-related symptoms: a randomized placebo-controlled, triple-blind aggregate n-of-1 clinical trial": Baseline demographics

Age: Total Group

| Age (years) | Number of Participants |
| --- | --- |
| 20-29 | 1 |
| 30-39 | 4 |
| 40-49 | 16 |
| 50-59 | 20 |
| 60-69 | 30 |
| 70-79 | 15 |
| 80-89 | 5 |

Primary Cancer Diagnosis: Total Group

Number of Participants

| Primary Cancer Diagnosis | Number of Participants |
| --- | --- |
| Blood/Lymph System | 13 |
| Brain | 1 |
| Breast | 34 |
| Gastrointestinal | 11 |
| Genitourinary | 8 |
| Gynecologic | 8 |
| Head/Neck | 5 |
| Lung | 2 |
| Sarcoma | 2 |
| Skin | 2 |
| Other | 4 |
| No personal history of cancer | 1 |

Cancer Stage: Total Group

Number of Participants

| Cancer Stage | Number of Participants |
| --- | --- |
| 0 (no evidence of disease) | 23 |
| I | 9 |
| II | 10 |
| III | 14 |
| IV | 28 |
| No personal history of cancer | 1 |
| Unknown | 6 |
