## Supplementary figures and images for "Comparing cannabinoid extracts for treating cancer-related symptoms: a randomized placebo-controlled, triple-blind aggregate n-of-1 clinical trial"

### PGIC and ESASrSN in same order

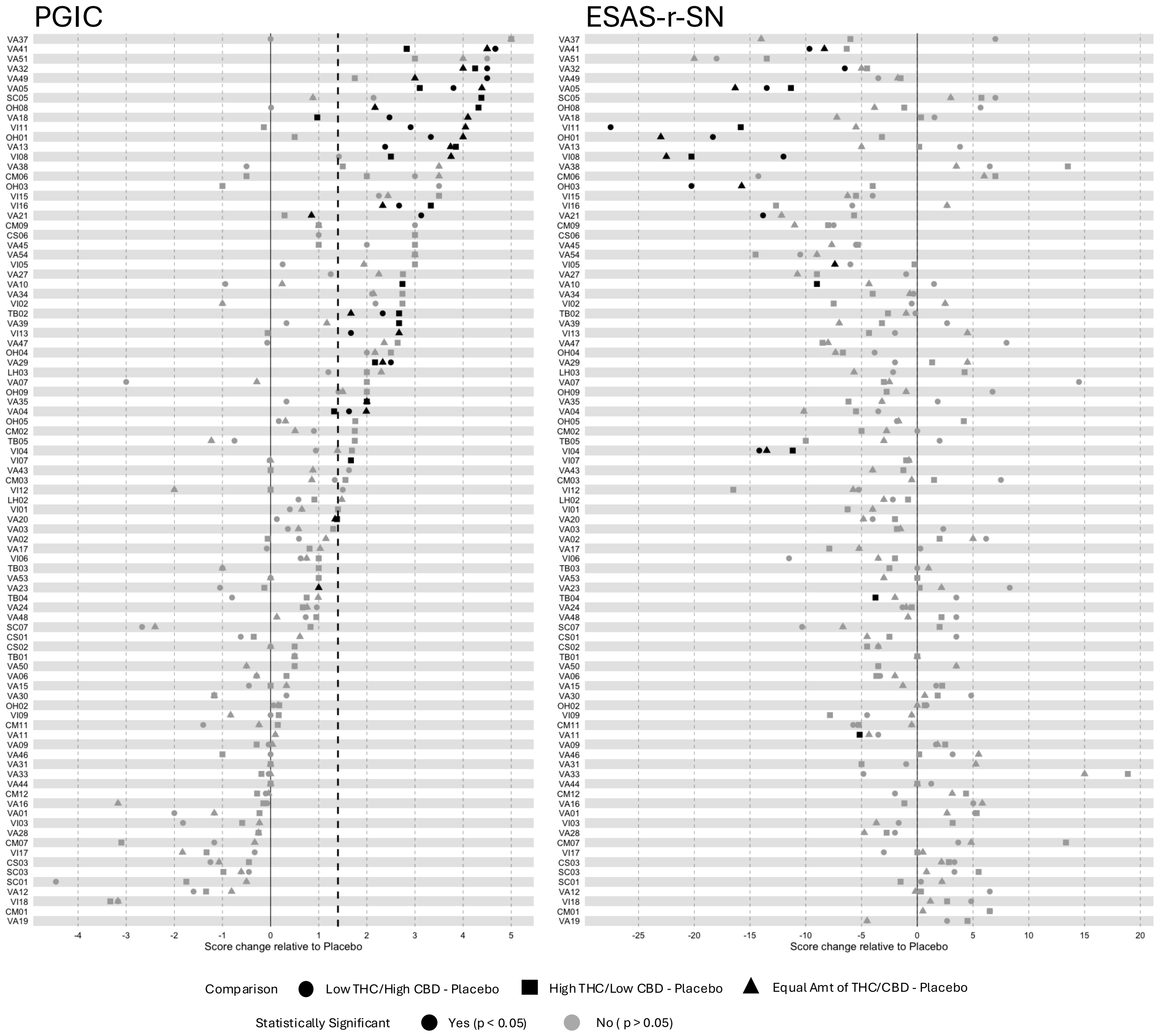
