## Supplementary material for "Comparing cannabinoid extracts for treating cancer-related symptoms: a randomized placebo-controlled, triple-blind aggregate n-of-1 clinical trial": Related adverse events

**Table 5. Logistic regression for risk of moderate/severe adverse events deemed possibly or probably related to each extract**

| Variable | Odds Ratio | 2.5% Confidence Interval | 97.5% Confidence Interval |
| --- | --- | --- | --- |
| (Placebo) | 0.022 | 0.009 | 0.045 |
| 1:1 | 1.061 | 0.390 | 3.148 |
| CBD | 0.918 | 0.302 | 2.881 |
| THC | 2.897 | 1.310 | 7.673 |
