## Supplementary material for "Comparing cannabinoid extracts for treating cancer-related symptoms: a randomized placebo-controlled, triple-blind aggregate n-of-1 clinical trial": Demographic logistic regression

| Predictor | Odds of response ratio | 95% Confidence intervals | p |
| --- | --- | --- | --- |
| Age at consent | 0.97 | 0.93-1.00 | 0.121 |
| Pain subgroup | Reference | | |
| Anxiety subgroup | 1.23 | 0.40-3.78 | 0.710 |
| Sleep subgroup | 1.74 | 0.57-5.47 | 0.336 |
| Female | Reference | | |
| Male | 0.75 | 0.24-2.34 | 0.611 |
| Stage 0 | Reference | | |
| I | 0.5 | 0.09-2.57 | 0.410 |
| II | 1.54 | 0.30-9.25 | 0.612 |
| III | 1.90 | 0.44-9.14 | 0.398 |
| IV | 0.88 | 0.25-3.09 | 0.945 |

Table 6. Demographics logistic regression (Obs=84, R^2^Tjur 0.0870)
